# γδ T cells are expanded in the blood of chronic low back pain patients with lumbar Modic changes

**DOI:** 10.64898/2026.11.10.26367714

**Authors:** Klara Mocova, Pamela Bitterli, Alexandre Meier, Luan Phelipe Hatt, Andrea Nüesch, Danilo Menghini, Mazda Farshad, Ladislav Senolt, Florian Brunner, Jiri Baloun, Oliver Distler, Stefan Dudli, Jan Devan

## Abstract

**Objectives:** Modic changes (MC) are vertebral endplate and bone marrow lesions strongly associated with chronic low back pain. Type 1 MC (MC1, oedema-like) and type 2 or 3 MC (MC2or3, fat- and sclerosis-like) are commonly regarded as different stages of the same pathological process, yet their underlying biology remains poorly defined. In particular, it remains unclear whether MC are confined to the spine or accompanied by distinct systemic immune alterations. Here, we provide the first comprehensive characterisation of the circulating immune landscape in patients with lumbar MC.

**Methods:** Peripheral blood mononuclear cells were analysed from 37 patients undergoing lumbar spinal surgery: 11 with no MC, 15 with MC1 and 11 with MC2or3. Samples were fluorescently barcoded, pooled, and stained with a 28-antibody phenotyping panel. Each pool was then split into 30 aliquots, and each of these was co-stained with a different PE-conjugated anti-integrin, -selectin, or -chemokine receptor antibody or isotype controls. Differences in abundance were tested with binomial mixed models adjusted for acquisition batch, age, and sex. Differentially abundant compartments were clustered by FlowSOM and the same binomial model was used to test differences in cluster abundance. Adhesion receptors were analysed as single chains.

**Results:** The γδ T cell compartment was the only population significantly expanded in MC patients (median 0.58% of all recorded cells in nonMC donors versus 1.61% in pooled MC (OR = 2.38, q = 0.035)). Clustering γδ T cells into subsets showed compartment-wide expansion, primarily driven by Vδ2^+^ T cells. The adhesion receptor screen revealed α6 and α2 integrin on basophils, endoglin on plasmacytoid dendritic cells, and β7 integrin on IgA⁺ B cells as differentially expressed in MC, but none survived multiple-testing correction across the whole screen.

**Conclusions:** Lumbar MC are accompanied by expansion of circulating γδ T cells. This aligns with previously reported γδ T cell accumulation in MC1 lesions, underscoring the role of γδ T cells in MC pathophysiology.

## Introduction

Low back pain is one of the leading global causes of years lived with disability^1^ and although most acute episodes resolve, roughly 20% of them progress to chronic low back pain^2^. Modic changes (MC) are vertebral endplate and bone marrow signal alterations visible on magnetic resonance imaging (MRI)^3^ present in 20–50% of patients with chronic low back pain, in contrast to a significantly smaller fraction of asymptomatic individuals^4,5^. They are classified into three types by their appearance on T1- and T2- weighted MRI: type 1 (MC1), with oedema-like signal and fibrovascular marrow; type 2 (MC2), with fatty marrow replacement; and type 3 (MC3), with sclerosis^3^. MC are an independent risk factor for episodes of severe and disabling pain, predict poorer clinical outcome, and patients with MC1 report an inflammatory pain pattern^6–10^. Despite this clinical importance, there is no consensus on how to treat patients with MC, and the absence of disease-modifying therapies reflects the limited understanding of MC pathobiology.

Two competing MC pathomechanistic explanations dominate. The first is autoinflammatory. MC occur almost exclusively adjacent to degenerate discs and damaged endplates. Endplate defects expose nucleus pulposus material to the immune cells in bone marrow, provoking a local inflammatory reaction with raised proinflammatory cytokines and neoinnervation of the endplate^11–16^. Animal work supports this notion: endplate damage exposes normally immune-privileged nucleus pulposus tissue to marrow immune cells and produces lesions resembling MC with significant T cell accumulation within it^17,18^. In humans, the disc and the adjacent marrow show inflammatory crosstalk^19^, with lesional marrow neutrophils actively degrading the cartilage endplate^20^. The second explanation is a low virulent disc infection. Bacteria, principally *Cutibacterium acnes*^21^, colonise the disc and drive a chronic marrow response^22,23^. This hypothesis underpinned subsequent antibiotic trials, whose outcomes remain controversial^24,25^. The common denominator of both hypotheses is immune system activation.

Accordingly, genetic data also implicate a significant role of the immune system in MC. MC are heritable (30% heritability), and polymorphisms of the IL1 gene and PTPRD gene represent known risk factors^26–28^. We also discovered that certain MHC alleles are associated with a significantly increased risk of MC development. HLA-A*26 carriers have a significantly increased probability of MC development^29^. For MC1 specifically, the HLA 8.1 ancestral haplotype has been associated with the formation of lesions that mediate clinically relevant chronic low back pain. This haplotype spans several regions. It covers class I and II genes encoding molecules that present peptides to conventional T cells, and the class III region encoding tumour necrosis factor and complement. Adjacent to HLA-B in the class I region, it also covers the stress-inducible NKG2D ligands MICA and MICB^30^. This is therefore a signal compatible with peptide antigen presentation as well as with innate or innate-like recognition of stressed cells^30,31^.

The systemic immune response in MC remains largely unexplored. Mechanistic studies have focused almost exclusively on disc and endplate tissue obtained at surgery, defining local immune alterations but not whether they extend into the circulation. The few blood-based studies have largely measured soluble inflammatory markers, with mostly negative or inconsistent findings^32–37^. Whole blood RNA sequencing in antibiotic trial patients correlated interferon signalling, mitochondrial metabolism, and antiviral defence genes with STIR lesion volume^38^. Interestingly, blood immune cell activation in MC subjects has recently been linked to intradiscal propionic acid^39^, which hints at a link between the immune system and the spine. Yet it is unknown whether specific immune cell subsets accumulate in the blood of MC patients, and whether they could explain the MC lesion-systemic link. Understanding the mechanisms underlying this link and the immune infiltration observed in MC could help uncover novel treatment targets or strategies to alleviate disease burden.

To address this gap, we performed the first comprehensive flow cytometry-based immunophenotyping of patients with lumbar MC, alongside a parallel screen of 28 integrin, selectin, and chemokine receptors. This strategy allowed us to simultaneously determine whether specific immune cell populations differ in frequency and whether they exhibit distinct trafficking phenotypes.

## Materials and Methods

### Study design, patients and samples

Patients undergoing elective lumbar spinal surgery were recruited at Balgrist University Hospital. MC status was assigned from preoperative T1- and T2-weighted MRI and recorded per lumbar level. Patients with known presence of diseases that influence the composition of the immune system were excluded (HIV, cancer, discitis, autoimmune diseases, acute infections). Patients were assigned to the MC group if an MC was present at any lumbar level, including levels not operated on, since a systemic readout cannot distinguish lesion location. Patients whose lesions included an MC1 component (MC1, MC1/2, MC1/3, MC1/2/3) were assigned to MC1; patients with MC without MC1 components (MC2, MC3, MC2/3) were assigned to MC2or3. The nonMC group comprised patients undergoing lumbar surgery with no MC at any level of the lumbar spine; four of them were operated on for extension of a previous fusion.

Venous blood was taken preoperatively. Blood was collected into tubes with EDTA and immediately processed. Peripheral blood mononuclear cells (PBMCs) were isolated using density gradient centrifugation on Ficoll, resuspended in heat-inactivated fetal calf serum supplemented with 10% dimethyl sulfoxide, and stored in liquid nitrogen. Visual analogue scale (VAS) back pain and Oswestry Disability Index (ODI) scores were obtained through questionnaires^40,41^.

The study was conducted in accordance with the Declaration of Helsinki and was approved by the Cantonal Ethics Committee of Zurich (KEK-ZH, BASEC 2018-01486). All patients gave written informed consent.

### Immunophenotyping and massively parallel flow cytometric screening of PBMCs

PBMCs were thawed and stained on ice, each sample with a unique combination of one to three anti-CD45 conjugates so that every sample in a batch carried its own fluorescent barcode^42^. After the initial staining, the samples of a batch were pooled and stained together with an identical 28-antibody phenotyping panel (**Tab. S1**). The pool was then split into 30 aliquots, 28 receiving one PE-conjugated antibody each against an integrin, selectin, or chemokine receptor and the remaining two a matched isotype control, together with a viability dye. Samples were acquired on a five-laser Aurora (Cytek) in two acquisition batches and deconvolved by barcode (**Tab. S1**). The detailed staining procedure is described in the Supplementary Methods.

### Gating

Twenty-six immune cell populations were analysed (**Fig. S1**). Gates were applied programmatically to all 1,080 donor × marker samples using CytoML, flowWorkspace and flowCore^43–45^. The gating tree, the

residues excluded from it and the isotype-based gates for the screened antibodies are described in the Supplementary Methods.

### Marker positivity thresholds

To identify cluster identity and analyse T cell functional maturation, we set positivity thresholds, turning each marker’s continuous signal into a positive or negative call for each cell. Rather than splitting each marker’s histogram, every threshold was anchored to the original gating tree, cutting where the F1 score peaks. Markers with no clean anchor keep a two-component Gaussian mixture cut (mclust)^46^. **Tab. S2** gives every threshold with its calibration, and Fig. S2 shows the z-scored mean signal of every backbone marker in all 26 gated populations. Supplementary Methods describe further details.

### Statistical analysis

#### Analysis of differential abundance of immune populations

For each population, we modelled the number of its cells out of all CD45-positive cells recorded in the sample. The model was a binomial mixed model with fixed effects for group, acquisition batch, age, and sex, and a random intercept for each patient^47^. The group effect was tested by likelihood ratio test against the same model without it. Three comparisons against nonMC samples were reported for every population: pooled MC, MC1, MC2or3. Confidence intervals were Wald intervals on the log odds ratio. Benjamini–Hochberg correction was applied once across all three comparisons and all 26 named populations together across 78 tests. Patient characteristics were compared across the three groups with the Kruskal–Wallis test and sex with Fisher’s exact test. The Supplementary Methods provide further details.

#### Subclustering of the selected cell subsets

The CD8 compartment, the γδ compartment, the Vδ2 subset within the γδ compartment, the basophil compartment and the B cell compartment were subclustered separately. Gate membership was taken from the applied workspace for each well. Measured backbone channels were asinh(x/1500) transformed as previously described^48^ and z-scored within the acquisition batch.

Only markers that could vary within the defining gate were included. The CD8 subclustering used 12 markers: CD7, CD11b, CD16, CD25, CD28, CD38, CD45RA, CD56, CD127, HLA-DR, NKG2D and PD-1. The γδ subclustering used 19 markers: CD4, CD7, CD8, CD11b, CD11c, CD16, CD25, CD28, CD38, CD45RA, CD56, CD127, CD161, HLA-DR, NKG2D, PD-1, pan-γδ TCR, TCR Vδ1 and TCR Vδ2. The Vδ2 subclustering used 16 markers: CD4, CD7, CD8, CD11b, CD11c, CD16, CD25, CD28, CD38, CD45RA, CD56, CD127, CD161, HLA-DR, NKG2D and PD-1. The basophil subclustering used 8 markers: CD4, CD7, CD11b, CD11c, CD25, CD28, CD45RA and CD127. The B cell subclustering used 7 markers: CD11c, CD24, CD25, CD38, CD45RA, IgA and IgD.

Clustering was performed using a 10 × 10 self-organising map implemented in FlowSOM^49,50^, an established approach for cytometry analysis^51^. The FlowSOM codebook was subsequently partitioned by k-means, weighted by the number of cells assigned to each map unit. This ensured that the partition was driven by cell mass rather than by sparsely populated or empty regions of the map. For visualisation, UMAP embeddings of the same scaled cells were generated using the uwot implementation^52^. UMAP was used for display only. No results were inferred from the embedding, and all abundance analyses were performed on the FlowSOM clusters. Cluster abundance was expressed as the proportion of all recorded cells and, where stated, of the parent compartment.

#### Selection of the number of clusters

K was chosen by bootstrap Jaccard cluster stability under a size constraint^53^. Final K is the largest value satisfying three rules at once: the size-weighted mean Jaccard is at least 0.70, no single cluster falls below 0.60, and no cluster holds less than 5% of the compartment. The second rule matters because a high mean can hide one unreproducible cluster among stable ones; the third prevents a nominally stable partition that isolates a fraction of a per cent of the cells.

#### Differentiation staging and the functional maturation axis

Cells were assigned to the CD8 differentiation sequence described by Larbi and Fulop^54^: N1 → M1 (central memory) → M2/M3 (effector memory) → M4 (TEMRA). Four markers from their panel were available in our dataset: CD45RA, CD28, PD-1, and CD127. Because M2 and M3 differ only in markers not included in our panel, they were combined into a single effector-memory stage.

The differentiation stage was assigned by gating rather than clustering. Each cell was classified using its CD45RA and CD28 status at the anchored positivity thresholds: N1 = CD45RA⁺CD28⁺, M1 = CD45RA⁻CD28⁺, M2/M3 = CD45RA⁻CD28⁻, M4 = CD45RA⁺CD28⁻. Stage proportions were tested using the same binomial mixed model described above.

To represent differentiation as a continuous axis, each cell was projected onto a smoothed path through the four stage centroids in the order described by Larbi and Fulop^54^. Arc-length along this path was rank- normalised to [0, 1]. Thus, the direction of the axis was specified a priori by the differentiation stages rather than inferred from the data, as in diffusion pseudotime^55^. This matters because a trajectory fitted freely across the entire γδ compartment primarily ordered cells according to δ chain expression.

Differences between groups along the continuous axis were tested without selecting a cut point on this axis. The pooled trajectory was divided into ten equal bins, and each donor’s cell counts across bins were analysed jointly in a single Poisson mixed model. Group, bin, and their interaction were included as fixed effects, together with batch, age, and sex. The log of each donor’s total number of cells on the axis was included as an offset, so the model tested differences in the shape of the distribution along this axis rather than differences in total cell number. The full model was compared with a model lacking the group × bin interaction using a likelihood ratio test, yielding a single omnibus *p-value* per comparison. Thus, no cut point was selected along the axis, and no multiple-testing correction across bins was required. An observation-level random effect was included to absorb overdispersion, so that the comparison is between donors rather than between cells^56^.

#### Adhesion receptor screen at the level of single chains

For each of the 28 PE-conjugated integrin, selectin, and chemokine receptor antibodies, we applied the same mixed model to PE-positive versus PE- negative counts within each population. Analyses were restricted to antibody × population combinations in which the marker was expressed, defined as ≥ 30 positive cells with a mean frequency ≥ 0.5%, or ≥ 10 cells and ≥ 2%, in at least one of the three groups (nonMC, MC1 or MC2or3). Pairs with fewer than four donors in a group after quality filtering were not modelled. Where the pooled PE distribution was not bimodal (αL integrin, β2 integrin and ICAM-1 in every population) or more than 85% of the cells were positive, the readout was instead the log₂ median PE intensity per donor, compared between groups with a linear model with the same fixed effects; these 154 pairs are marked “median intensity”, and their effects are differences in log₂ median intensity. P values were corrected within each population (families of at most 28 tests) and separately across the whole screen (Supplementary Methods). The expression map shows, for each antibody × population pair, the percentage of PE-positive cells pooled across all donor wells minus the percentage of the matched isotype control above the same gate, with negative values set to 0. Populations and antibodies were ordered by hierarchical clustering of this map (Euclidean distance on log(1 + %) values, average linkage), and the same order was used for the effect map. The number of nominally significant pairs was calibrated by permutation: the MC label was shuffled between donors and the whole screen refitted 83 times (Supplementary Methods).

#### Reporting statistical significance

Results that remained statistically significant after multiple-testing correction are reported with their q- value. Results with nominal *p* < 0.05 that did not remain significant after correction are reported with their unadjusted *p*-value and described as nominally significant. The full statistical output is provided as Supplementary Data 1.

## Results

### Patient characteristics

Thirty-seven patients were analysed (11 nonMC, 15 MC1, and 11 MC2or3; **Fig. 1A**). Visual analogue scale (VAS) back pain and Oswestry Disability Index (ODI) scores were available for 36 and 35 of the 37 patients, respectively^40,41^. The groups did not differ in age, sex, or VAS back pain (**Fig. 1B, C**), but differed in ODI (median 54, 53, and 30, respectively; p = 0.009) (**Fig. 1C** **right**).

**Fig. 1.**
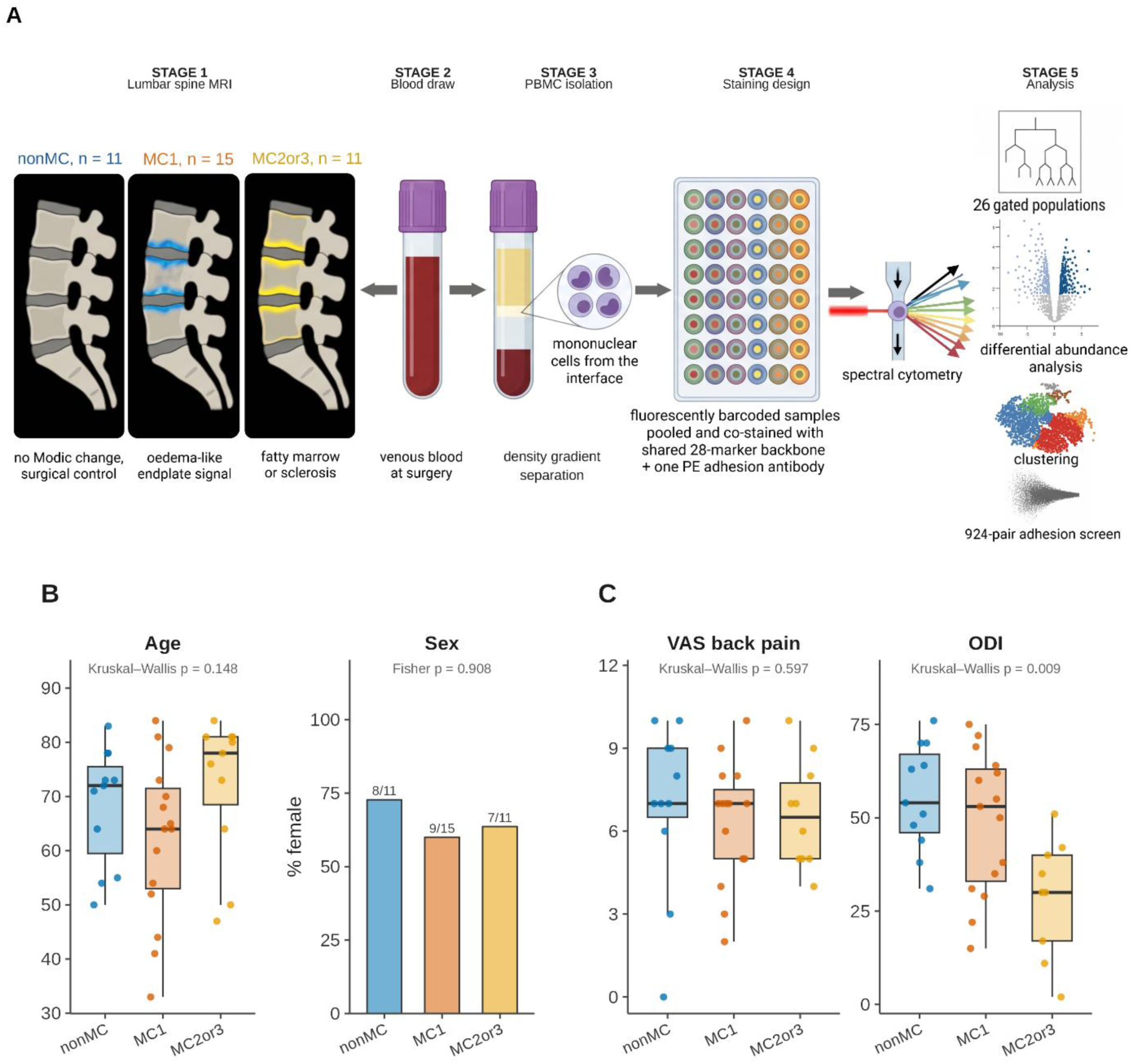
Cohort description and study design. (**A**) Schematic representation of study design. (**B**) Age and sex distribution by group. (**C**) VAS back pain and ODI differences between the groups; boxes are median and interquartile range, whiskers are 1.5 × that range. Kruskal–Wallis test for age, VAS and ODI; Fisher’s exact test for sex.

### MC are accompanied by a γδ T cell expansion

Each population was compared against nonMC in three comparisons: MC1 vs nonMC, MC2or3 vs nonMC, and both MC groups pooled vs nonMC. γδ T cells were the only population significantly increased in donors with MC when both MC groups were pooled (pooled MC OR = 2.38, q = 0.035) (**Fig. 2A, B, C**), with the highest median frequency in MC1 (nonMC: 0.58%, MC1: 2.34%, MC2or3: 1.48%). Among γδ T cell subsets, Vδ2 cells showed the strongest increase (pooled MC OR = 2.62, p = 0.007), whereas Vδ1⁺ (pooled MC OR = 1.83, p = 0.087) and Vδx (γδ T cells expressing neither Vδ1 nor Vδ2; pooled MC OR = 1.73, p = 0.071) showed similar but non-significant increases (**Fig. 2C**).

**Fig. 2.**
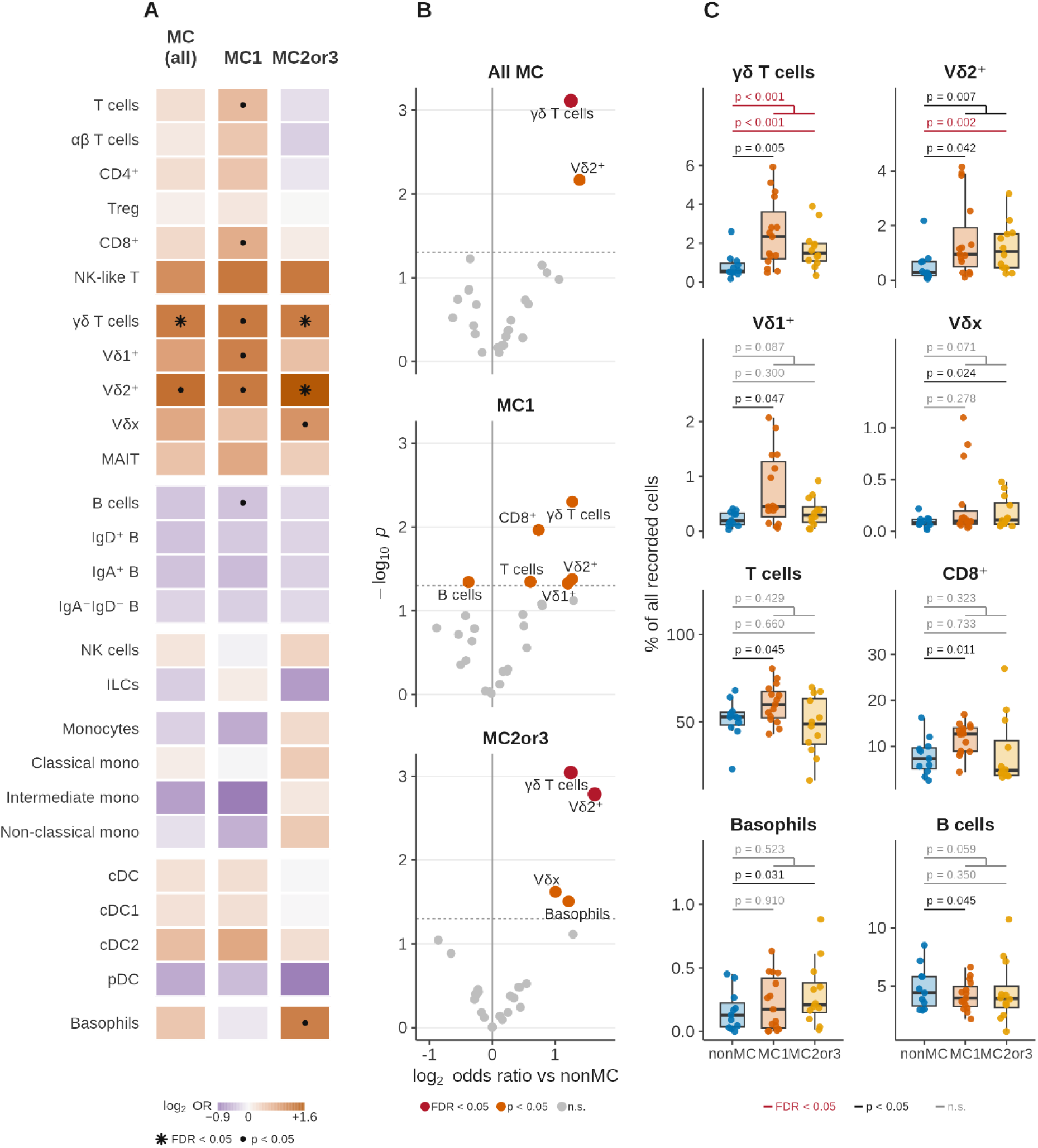
**Expansion of γδ T cells in MC patients’ blood**. All 26 gated populations as percentages of all recorded cells, each tested against the nonMC group with a binomial mixed model adjusted for batch, age, and sex. (**A**) log₂ odds ratios in three comparisons, pooled MC (All MC or MC (all) in the figures), MC1 and MC2or3; orange higher in MC, purple lower; ✱ survives correction, ● nominally significant; rows follow the gating hierarchy in lineage blocks. (**B)** The same comparisons as volcano plots; red survives correction, orange nominal, grey neither; dashed line p = 0.05. (**C**) The eight populations carrying the signal. The three brackets give p for MC1, MC2or3 and pooled MC against nonMC, from bottom to top. Red: p-value surviving correction; black: nominal; grey: neither.

Comparing MC1 with nonMC, several differences reached nominal significance, but none survived multiple-testing correction. γδ T cells were increased to a similar extent as in MC2or3 (OR = 2.42, p = 0.005). Both Vδ1⁺ (OR = 2.30, p = 0.047) and Vδ2⁺ (OR = 2.41, p = 0.042) were also nominally increased. In addition, MC1 showed nominal changes in conventional T cells that were not observed in the pooled MC comparison: CD8⁺ αβ T cells (hereafter CD8 T cells; OR = 1.67, p = 0.011) and total T cells (OR = 1.52, p = 0.045) were increased, accompanied by a decrease in B cells (OR = 0.77, p = 0.045). Interestingly, none of the three populations differed significantly between MC2or3 and nonMC (**Fig. 2A, B**).

In the MC2or3 vs nonMC comparison, γδ T cells (OR = 2.38, q = 0.035) and Vδ2⁺ cells (OR = 3.10, q = 0.043) remained significantly increased after multiple-testing correction. Vδx T cells were nominally increased (OR = 2.01, p = 0.024), whereas Vδ1⁺ showed no significant difference despite an OR in the same direction (OR = 1.46, p = 0.300). Basophils were also nominally increased (OR = 2.32, p = 0.031) (**Fig. 2A, B**).

### Inside the γδ T cell compartment: all subsets and all functional maturation stages expand

To investigate whether the observed accumulation of γδ T cells in the blood of patients with MC reflects expansion of specific subsets, the 135,845 γδ T cells that entered the subclustering from the 37 patients were clustered using FlowSOM based on the 19 markers retained for the γδ T cell subclustering (**Fig. 3**). Four clusters were retained (K = 4, mean J = 0.86, weakest cluster J = 0.77; **Fig. S3A to C**). The clusters

**Fig. 3.**
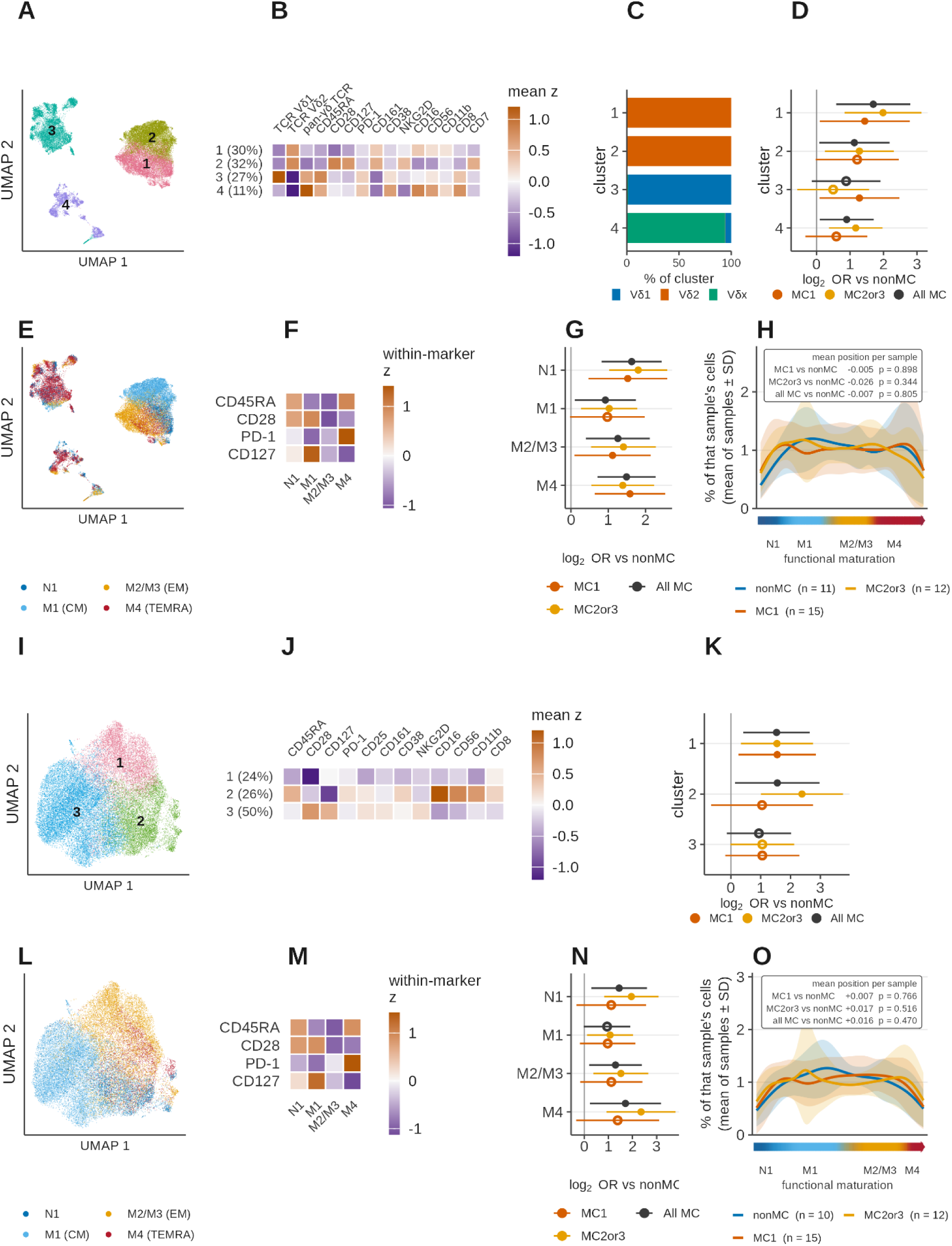
**γδ cell clustering**. Rows one and two: the γδ compartment; rows three and four: the Vδ2 subset. (**A**) UMAP by FlowSOM showing the (1) cytotoxic Vδ2⁺, (2) resting Vδ2⁺, (3) Vδ1⁺, and (4) Vδx clusters. (**B**) Cluster phenotypes for the markers that separated the clusters. (**C**) δ chain composition of each cluster. (**D**) Cluster abundance as a share of all recorded cells; filled circles, p < 0.05; open circles, not significant. (**E**) UMAP by functional maturation stage. (**F**) Functional maturation stage phenotypes. (**G**) Functional maturation stage abundance, as in (D). (**H**) Distribution of each sample’s cells along the functional maturation axis from N1 to M4; lines are group means of the per-sample densities, shaded ± SD across samples. The inset gives the shift in mean position per sample with its p-value; the text reports the omnibus test across bins (Methods). (**I**) UMAP by FlowSOM showing the (1) CD28⁻CD127⁺, (2) CD16⁺CD56⁺CD11b⁺ cytotoxic, (3) CD28⁺CD127⁺CD161⁺ resting clusters of Vδ2^+^ T cells. (**J**) Cluster phenotypes for the markers that separated the clusters. (**K**) Cluster abundance, as in (D). (**L**) UMAP by functional maturation stage. (**M**) Functional maturation stage phenotypes. (**N**) Functional maturation stage abundance, as in (D). (**O**) Distribution along the functional maturation axis from N1 to M4; group means, shaded ± SD across samples; inset as in (H).

were distinguished primarily by the δ-chain and secondarily by differentiation or activation markers (**Fig. 3A to C**). They comprised a cytotoxic Vδ2⁺ cluster (29.5%; CD16 53%, CD56 52%, CD11b 53%, CD28 23%), a resting Vδ2⁺ cluster (31.8% of γδ T cells; CD28 85%, CD127 96%, CD161 39%), a Vδ1⁺ cluster (27.5%; CD45RA 85%, CD127 83%, CD56 43%), and a Vδx cluster (11.2%; CD11c 21%, HLA-DR 17%, CD4 26%, CD38 35%, PD-1 25%).

When expressed as share of all recorded cells, all four γδ T cell clusters were higher in the pooled MC group (**Fig. 3D**): cytotoxic Vδ2⁺ cluster (pooled MC: OR = 3.24, q = 0.017; MC2or3: OR = 3.96, q = 0.008); resting Vδ2⁺ cluster (pooled MC: OR = 2.19, p = 0.040; MC2or3: OR = 2.43, q = 0.028), Vδx cluster (pooled MC: OR = 1.87, p = 0.034; MC2or3: OR = 2.25, q = 0.016). The Vδ1⁺ cluster reached nominal significance only in the MC1 comparison (OR = 2.43, p = 0.042). In contrast, when cluster abundance was expressed relative to the γδ T cell compartment, none of the four clusters changed significantly. Thus, the expansion of circulating γδ T cells reflects expansion of the compartment as a whole rather than selective expansion of a single subset.

The differentiation analysis led to the same conclusion. Each γδ T cell was assigned to one of the four functional maturation stages based on its CD45RA and CD28 expression. N1 (CD45RA⁺CD28⁺), M1 central memory (CD45RA⁻CD28⁺), M2/M3 effector memory (CD45RA⁻CD28⁻), M4 TEMRA (CD45RA⁺CD28⁻). As expected, PD-1^+^ cells were most frequent in the TEMRA stage, whereas CD127 was highest in central memory cells and lowest in effector memory cells (**Fig. 3E, F**). When expressed as a proportion of the γδ T cell compartment, none of the stages differed significantly after multiple-testing correction. N1 was nominally increased in MC2or3 (OR = 1.66, p = 0.028) and showed a similar trend in pooled MC (OR = 1.43, p = 0.093). In contrast, when expressed as a proportion of all recorded cells, all four stages were increased and remained significant after correction in the pooled MC comparison: N1 (OR = 3.09, q = 0.001), M1 (OR = 1.89, q = 0.033), M2/M3 (OR = 2.39, q = 0.008), and M4 TEMRA (OR = 2.81, q = 0.001). Similar results were observed when MC1 and MC2or3 were compared separately with nonMC (**Fig. 3G**).

Consistent with these findings, the overall distribution of cells along the functional maturation axis was unchanged (MC1 p = 0.756, MC2or3 p = 0.682, pooled MC p = 0.639) (**Fig. 3H**). Together, these results indicate expansion of the γδ T cell compartment without a major shift in its differentiation.

### Vδ2⁺ T cell expansion spans phenotypic and differentiation states

Vδ2⁺ T cells are the largest subset in the γδ T cell compartment and showed the strongest increase in the gated analysis. We therefore analysed them separately using FlowSOM, clustering 83,288 Vδ2⁺ cells from 37 aliquot sets (36 patients; one nonMC set with fewer than 10 Vδ2⁺ cells was not subclustered) based on the 16 markers retained for the Vδ2 subclustering. Three clusters were retained (K = 3, mean J = 0.85, weakest cluster J = 0.81; **Fig. S3A to C**): a CD28⁺CD127⁺CD161⁺ resting cluster (49.8%), a CD16⁺CD56⁺CD11b⁺ cytotoxic cluster (25.8%, CD16 84%, CD56 73%, CD11b 74%, CD45RA 50%), and a CD28⁻CD127⁺ cluster (24.4%, CD28 4%, CD127 87%). The clusters and their phenotypes are shown in **Fig. 3I, J**.

When expressed as a proportion of all recorded cells, the CD28⁻CD127⁺ cluster was increased in all three comparisons, two of which survived multiple-testing correction (pooled MC: OR = 2.89, q = 0.030; MC1: OR = 2.93, p = 0.026; MC2or3: OR = 2.92, q = 0.027). The cytotoxic cluster was nominally increased in pooled MC (OR = 2.94, p = 0.037) and significantly in MC2or3 (OR = 5.22, q = 0.007), whereas the resting cluster showed increases in the same direction without reaching significance (**Fig. 3K**). In contrast, none of the three clusters changed significantly when expressed as a proportion of the Vδ2 compartment.

Differentiation staging divided Vδ2^+^ T cells into N1 (15.0%), central memory (40.2%), effector memory (33.3%), and TEMRA (11.6%), with monotonic functional maturation (**Fig. 3L, M**). Compared with the γδ T cell compartment overall, Vδ2⁺ T cells were more strongly represented in the memory stages. No differentiation stage changed significantly as a proportion of the Vδ2 T cell compartment. When expressed as a proportion of all recorded cells, however, N1, effector-memory, and TEMRA cells were increased in both pooled MC (N1 OR = 2.72, q = 0.037; effector memory OR = 2.45, q = 0.037; TEMRA OR = 3.26, q = 0.037) and MC2or3 (N1 OR = 3.87, q = 0.005; effector memory OR = 2.87, q = 0.020; TEMRA OR = 5.11, q = 0.007). Central memory Vδ2^+^ T cells were significantly increased in MC2or3 (OR = 2.10, q = 0.039), but not in the pooled MC (OR = 1.93, p = 0.062) (**Fig. 3N**). None of the stages differed significantly in MC1 (**Fig. 3N**). Consistent with these results, the distribution of Vδ2⁺ cells along the functional maturation axis did not differ from nonMC in any comparison (MC1 p = 0.858, MC2or3 p = 0.774, pooled MC p = 0.822) (**Fig. 3O**). The stage composition of the γδ, Vδ2, and CD8 compartments is compared in **Fig. S3D**.

Thus, as for the γδ T cell compartment overall, the increase in Vδ2⁺ T cells is distributed across phenotypic and differentiation states rather than being restricted to a single subset.

### CD8^+^ T cell changes in MC1 are concentrated in the naive subset

The CD8 αβ compartment was nominally increased in MC1 (**Fig. 2**). We therefore analysed 465,451 CD8^+^ T cells from all 37 patients using FlowSOM clustering based on the 12 measured markers retained for the CD8 subclustering (**Fig. 4**). Four clusters were identified (K = 4, mean J = 0.77, weakest cluster J = 0.71; **Fig. S3A to C**): a CD45RA⁻CD28⁺CD127⁺ NKG2D⁺ resting memory cluster (31.6%), an NK-like effector cluster (15.9%, CD56 85%, CD16 46%, CD11b 52%, CD28 20%), a PD-1⁺ cluster (18.7%, PD-1 75%, CD28 51%), and a CD45RA⁺ cluster (33.8%, CD45RA 93%, CD28 41%, CD127 89%) (**Fig. 4A, B**).

**Fig. 4.**
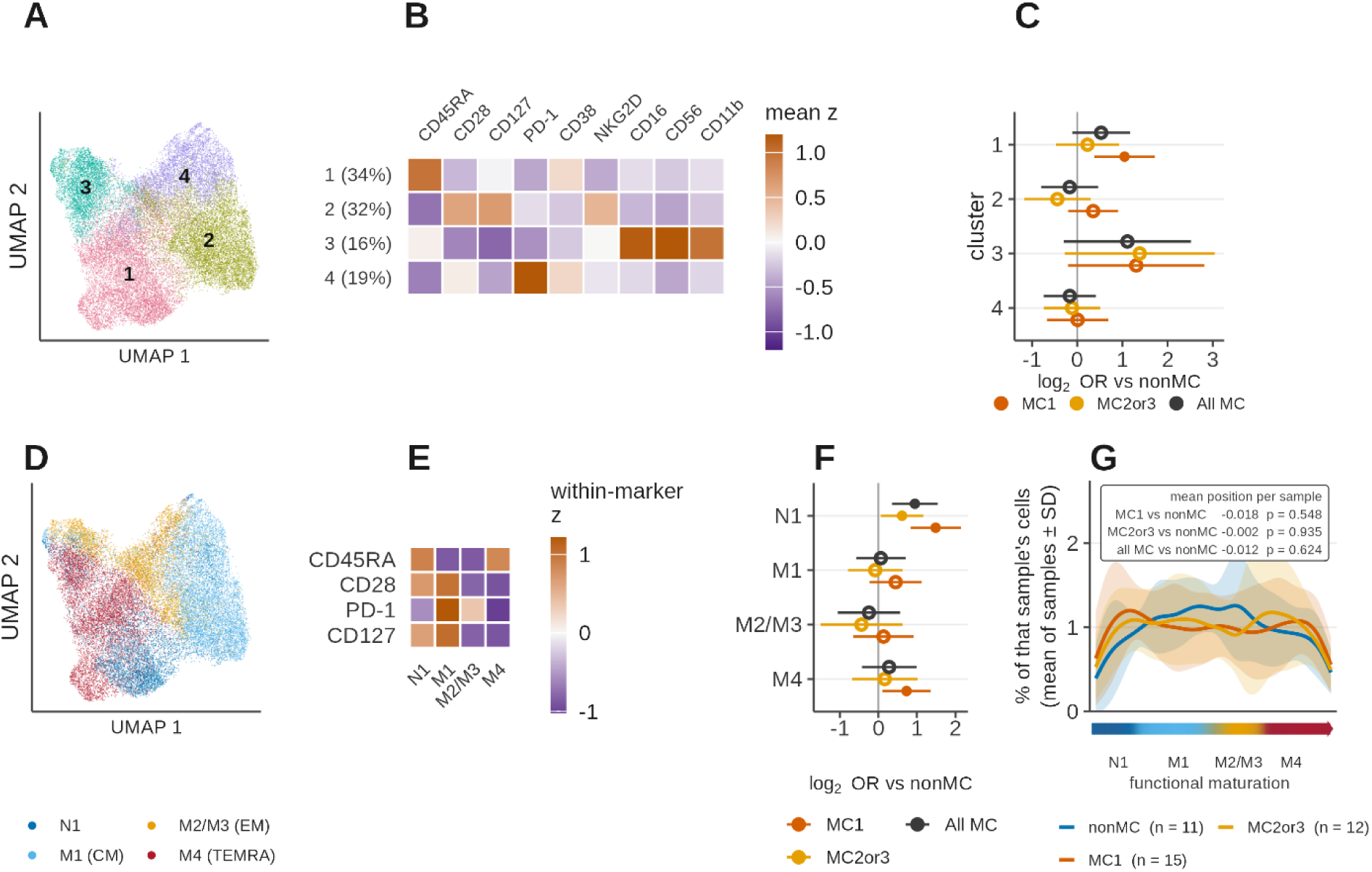
**CD8 T cell clustering**. (**A**) UMAP by FlowSOM showing the (1) CD45RA⁺, (2) CD45RA⁻CD28⁺CD127⁺ NKG2D⁺ resting memory, (3) NK-like effector, and (4) PD-1⁺ clusters. (**B**) Cluster phenotypes for the markers that separated the clusters. (**C**) Cluster abundance as a share of all recorded cells; filled circles, p < 0.05; open circles, not significant. (**D**) UMAP by functional maturation stage. (**E**) Functional maturation stage phenotypes. (**F**) Functional maturation stage abundance, as in (C). (**G**) Distribution of each sample’s cells along the functional maturation axis from N1 to M4; group means, shaded ± SD across samples; inset as in Fig. 3H.

In contrast to the γδ compartment, the composition of the CD8 compartment did change. The resting memory cluster was reduced as a proportion of CD8⁺ T cells in MC2or3 (OR = 0.60, p = 0.032) and in pooled MC (OR = 0.65, p = 0.033), while the PD-1⁺ cluster was reduced in MC1 (OR = 0.57, q = 0.034). When expressed as a proportion of all recorded cells, the CD45RA⁺ cluster was increased in MC1 (OR = 2.07, q = 0.018) (**Fig. 4C**).

Differentiation staging divided the CD8 compartment into N1 (20.4%), M1 central memory (31.1%), M2/M3 effector memory (19.3%), and M4 TEMRA (29.1%) (Fig. 4D, E). The clearest change involved the N1 naive stage. Its proportion within the CD8 compartment increased from 14.7% in nonMC to 20.1% in MC1 and 18.9% in MC2or3 (MC1: OR = 1.97, q = 0.015, MC2or3: OR = 1.65, p = 0.037; pooled MC: OR = 1.82, q = 0.010). N1 cells were also increased as a proportion of all recorded cells, from 0.74% in nonMC to 2.22% in MC1 and 1.31% in MC2or3 (pooled MC OR = 1.93, q = 0.012). TEMRA cells were nominally increased in MC1 only (OR = 1.66, p = 0.029). Central memory showed no significant change, while effector memory proportion was reduced without reaching significance (pooled MC OR = 0.64, p = 0.059) (**Fig. 4F**).

The overall distribution along the functional maturation axis did not differ significantly from nonMC in any comparison (MC1 p = 0.157, MC2or3 p = 0.241, pooled MC p = 0.096) (**Fig. 4G**). Thus, unlike the broad expansion observed in the γδ T cell compartment, the CD8 T cell compartment shows a more selective shift towards naive cells.

### Basophil and B cell changes are not subset-specific

The two remaining compartments that reached nominal significance in the gated analysis were examined using the same approach. For basophils, 19,267 cells from 30 aliquot sets (29 patients; eight sets with fewer than 30 basophils were not subclustered) formed three clusters (mean Jaccard 0.87, weakest cluster 0.72), comprising 13%, 34%, and 53% of the compartment (**Fig. 5A, B**). When expressed as a proportion of all recorded cells, all three clusters increased in MC, with the largest increase in cluster 1 (pooled MC OR = 2.78, p = 0.015) (**Fig. 5C**). In contrast, none of the clusters changed significantly as a proportion of the basophil compartment (smallest p = 0.080). Thus, the basophil increase was distributed across the compartment rather than restricted to a specific phenotype.

**Fig. 5.**
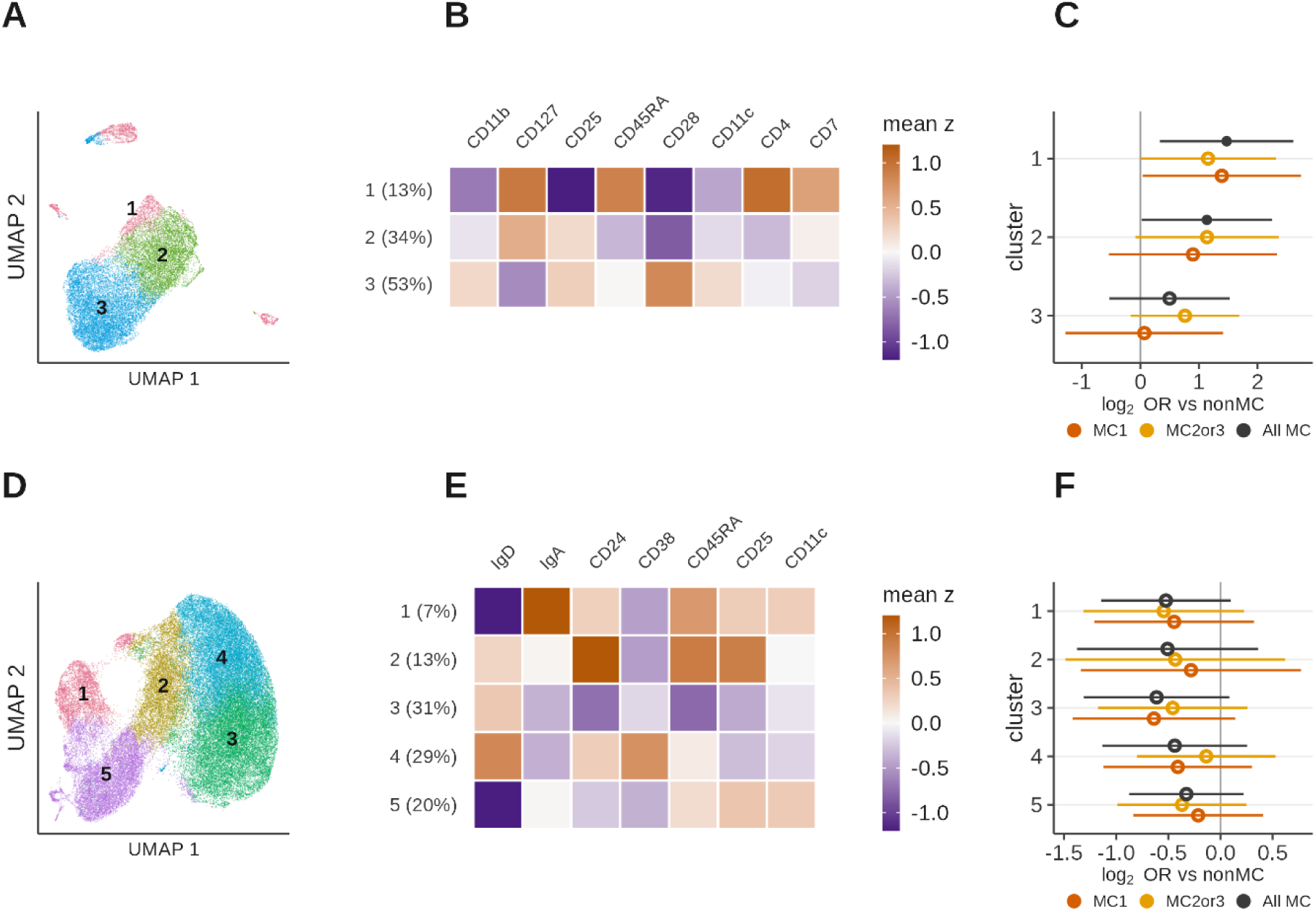
Clustering of the basophil and B cell compartments. (**A**) Basophil UMAP by FlowSOM cluster. (**B**) Basophil cluster phenotypes. (**C**) Basophil cluster abundance as a share of all recorded cells; filled circles, p < 0.05; open circles, not significant. (**D**) UMAP by FlowSOM cluster showing the (1) IgA⁺, (2) CD24^high^ IgD⁺, (3) mature naive, (4) CD38^high^ IgD⁺, and (5) IgD⁻IgA⁻ B cell clusters. (**E**) B cell cluster phenotypes. (**F**) B cell cluster abundance, as in (C).

For B cells, 309,855 gated events were available, of which 215,230 entered the subclustering and formed five clusters (**Fig. 5D, E**): mature naive cells (IgD⁺CD24^low^, 31%), two additional IgD⁺ clusters, characterised by CD24^high^ (13%) or CD38^high^ (29%), IgA⁺ class-switched memory (7%), and IgD⁻IgA⁻class-switched memory (20%). No individual B cell cluster differed significantly in abundance in any MC comparison, and none changed significantly as a proportion of the B cell compartment (**Fig. 5F**).

### The adhesion receptor screen yields candidates within populations

Twenty-eight integrin, selectin, and chemokine receptor antibodies were screened across immune populations. Expression above the matched isotype control (**Fig. 6A**) fell into three groups. β2, β1, α4, α5, α6 and αL integrin and ICAM-1 were present on nearly every population (median across populations 57% to 98% of cells). L-selectin, β7, β3 and α2 integrin and endoglin varied with the population, from below 1% to above 50%. The remaining 16 receptors were low or absent (median below 5%); αVβ6, α8, β4 and β5 integrin, CCR3 and CCR4 did not exceed 3% in any population (the chemokine receptor stainings are not interpretable, see Limitations). Basophils, classical monocytes and conventional dendritic cells expressed the largest number of receptors (13 to 15 at or above 10% of cells). After quality filtering, 673 antibody × population combinations were testable, of which 607 yielded fitted effects (**Fig. 6A, B**). Fourteen combinations remained significant after correction within the respective population. Among the recurrent findings, IgA⁺ B cells showed increased β7 integrin in pooled MC and MC1, while plasmacytoid dendritic cells showed reduced endoglin in pooled MC and MC2or3 and reduced L-selectin in MC2or3. Basophils showed increased α6 integrin in pooled MC and MC1 and increased α2 integrin in pooled MC. Several additional population-specific associations were observed (**Fig. 6B**). Every antibody × population pair in all three comparisons is listed in **Tab. S3**.

**Fig. 6.**
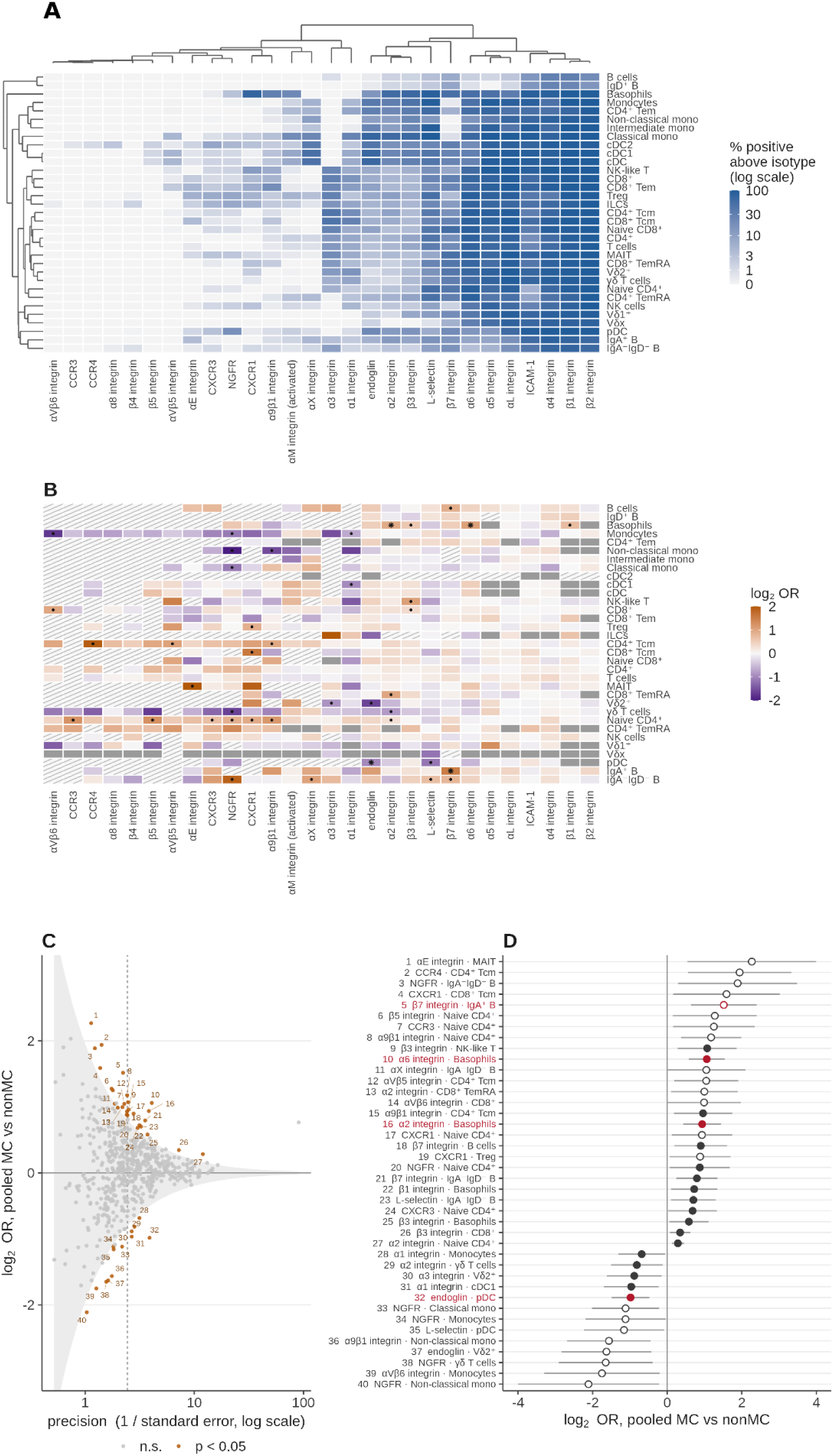
**Adhesion receptor screen**. (**A**) Pooled percentage of PE-positive cells per antibody × population over all donor wells, minus the matched isotype control; logarithmic blue scale; rows and columns ordered by hierarchical clustering. (**B**) Pooled MC comparison per testable pair as log₂ effect (odds ratio, or difference in log₂ median intensity for the 154 intensity readouts, same order as A; orange, higher in MC, purple, lower; ● nominally significant, ✱ surviving Benjamini–Hochberg correction within its population; hatched, marker not expressed on that population (251 pairs); grey, fewer than four donors left in a group – not tested. (**C**) Effect against precision for the 607 fitted pairs; nominally significant pairs numbered; grey band, chance at each precision; dashed line, median precision. (**D**) The 40 nominally significant pairs ranked by effect with 95% intervals; filled, better measured than the median pair; red, surviving correction within its population.

However, none of the associations remained significant after correction across the complete screen. Nominal associations were common (40 in pooled MC, 46 in MC1, and 53 in MC2or3; **Fig. 6C, D**), but permutation analysis of the pooled MC comparison showed that their number was compatible with chance: shuffled group labels produced a mean of 48.7 nominally significant associations, and the observed 40 were equalled or exceeded in 46 of 83 permutations (permutation p = 0.56). Thus, the adhesion-receptor screen identifies candidates for independent validation but provides no evidence for a screen-wide alteration in immune-cell trafficking receptors in MC. Analysis of the same markers across total PBMCs likewise showed no differences between groups (**Fig. S4A, B**).

## Discussion

Lumbar MC have been studied almost exclusively as a local process of the vertebral endplate and adjacent marrow. To our knowledge, this is the first comprehensive immunophenotyping of peripheral blood in patients with MC, showing that MC are accompanied by a measurable change in the composition of the circulating immune system. Importantly, this change is not diffuse, but is concentrated within the γδ T cell compartment, providing both a candidate blood-accessible readout of MC and a specific mechanistic lead. Vδ2⁺ cells, the largest of the three γδ T cell subsets, accounted for most of the observed increase and were the only subset consistently increased across all comparisons. Vδ1⁺ and Vδx cells showed changes in the same direction, but neither survived multiple-testing correction.

Vδ2⁺ T cells are the dominant circulating human γδ T cell subset^57,58^. They respond to phosphoantigens, small nonpeptide metabolites of isoprenoid biosynthesis^59^, sensed through the butyrophilins BTN3A1 and BTN2A1^60–63^. Microbial phosphoantigens, particularly (*E*)-4-hydroxy-3-methyl-but-2-enyl pyrophosphate from the bacterial nonmevalonate pathway, are several orders of magnitude more potent than the endogenous human ligand^64,65^ and persistent circulating Vδ2⁺ T cell expansion has been documented after several bacterial infections, including tularemia and Q fever^66,67^. However, Vδ2⁺ T cell activation is not specific to microbial exposure. Endogenous phosphoantigens can accumulate when the mevalonate pathway is perturbed in stressed or transformed cells. In addition, a small but reproducibly detectable population of circulating Vδ2⁺ T cells can recognise the Major Histocompatibility Complex (MHC) class I-related molecule 1 (MR1) loaded with endogenous metabolites, in an antigen-dependent but butyrophilin-independent manner^68^. Thus, Vδ2⁺ T cells can respond to both microbial and host-derived metabolic cues through TCR-dependent pathways. Such expansion is consistent with either the low- virulence bacterial hypothesis for MC^23,69^ or a sterile response to stress and tissue damage. In the latter settings, endogenous phosphoantigen can accumulate in metabolically stressed cells, MR1 can present self-metabolites to Vδ2⁺ T cells, and these cells can additionally respond to stress-induced NKG2D ligands on damaged cells^30,31,60^. Such signals are plausible in the inflamed and mechanically injured endplate and adjacent marrow in MC^12,14^. Our data cannot distinguish between microbial and sterile activation as the trigger of circulating Vδ2⁺ T-cell expansion.

The adhesion receptor screen provided no screen-wide evidence of altered immune-cell trafficking. Several population-specific candidates emerged, including increased β7 integrin on IgA⁺ B cells and reduced endoglin and L-selectin on plasmacytoid dendritic cells, but none survived correction across the full screen.

Previous attempts to identify a systemic signal in MC have focused predominantly on soluble markers, with the largest targeted and untargeted studies reporting no clear associations with MC^33,37^. Our findings are more closely aligned with studies identifying cell-associated systemic changes: first, an interferon and antiviral transcriptional program associated with MC lesion volume^38^, and second, blood immune cell activation associated with intradiscal propionic acid in MC1^39^.

In conclusion, lumbar MC are accompanied by an expansion of circulating γδ T cells, with the strongest signal in Vδ2⁺ T cells and in MC2or3. The expansion spans phenotypic and differentiation states, arguing against selective enrichment of a single γδ T-cell state. The differences between MC types may provide insight into the potential drivers of their divergent pathophysiology. Whether the observed γδ T-cell expansion reflects chronic microbial phosphoantigen exposure, sterile tissue stress, or another stimulus remains to be resolved. Nevertheless, these findings establish a systemic immune correlate of MC, broadening our understanding of the condition, which has long been considered primarily local to the affected vertebral tissues.

## Limitations

1. The cohort is small and does not provide sufficient power to discover changes with small effect size.
2. The “nonMC control” group is not healthy. These are lumbar surgical patients with comparable pain, four of whom were operated on for extension of a previous fusion. That makes the comparison conservative for detecting disease-associated differences, but heterogeneous.
3. The groups are matched on age, sex, and pain but not on disability: median ODI is 54 in nonMC and 53 in MC1 against 30 in MC2or3. Every comparison involving MC2or3 therefore carries that imbalance.
4. Acquisition batch coincides with collection period. It was handled by within-batch standardisation and as a model covariate rather than by a bridging control, which is the design we would recommend for replication.
5. MC type was assigned from clinical MRI without volumetric quantification, and mixed lesions were assigned to MC1 whenever a type 1 component was present. That is the conservative choice for detecting an MC1-specific signal, but it blurs the two groups.
6. Cytomegalovirus serostatus was not assayed, and CMV carriage is one of the strongest known determinants of the circulating γδ compartment^70,71^. It inflates the Vδ2-negative γδ cell subsets in particular^72^, which is not where our effect sits, but an imbalance between groups cannot be excluded.
7. MC were only examined at the lumbar spine, though they can also rarely occur in higher spine segments. MRI of the spine cranial to the lumbar region was not available.
8. Chemokine receptors are lost from the cell surface during cryopreservation and need time at 37 °C to be re-expressed. Samples were stained on ice immediately after thawing, which is optimal for immunophenotyping but does not allow for chemokine receptor re-expression. Therefore, staining for the four chemokine receptor antibodies (CCR3, CCR4, CXCR1, and CXCR3) was suboptimal.

## Data availability

The full statistical output for every comparison is provided as Supplementary Data 1, a workbook with one sheet per test family. FCS files and analysis scripts are available from the corresponding authors upon reasonable request.

## Supplementary Material

The Supplementary Material for this article comprises the Supplementary text, **Fig. S1** to **Fig. S4**, **Tab. S1** to **Tab. S3**, and Supplementary Data 1, the complete statistical output, one sheet per test family.

## Data Availability

All data produced in the present study are available upon reasonable request to the authors

