## Supplementary figures and images for "γδ T cells are expanded in the blood of chronic low back pain patients with lumbar Modic changes"

### Supplemental Figure 1

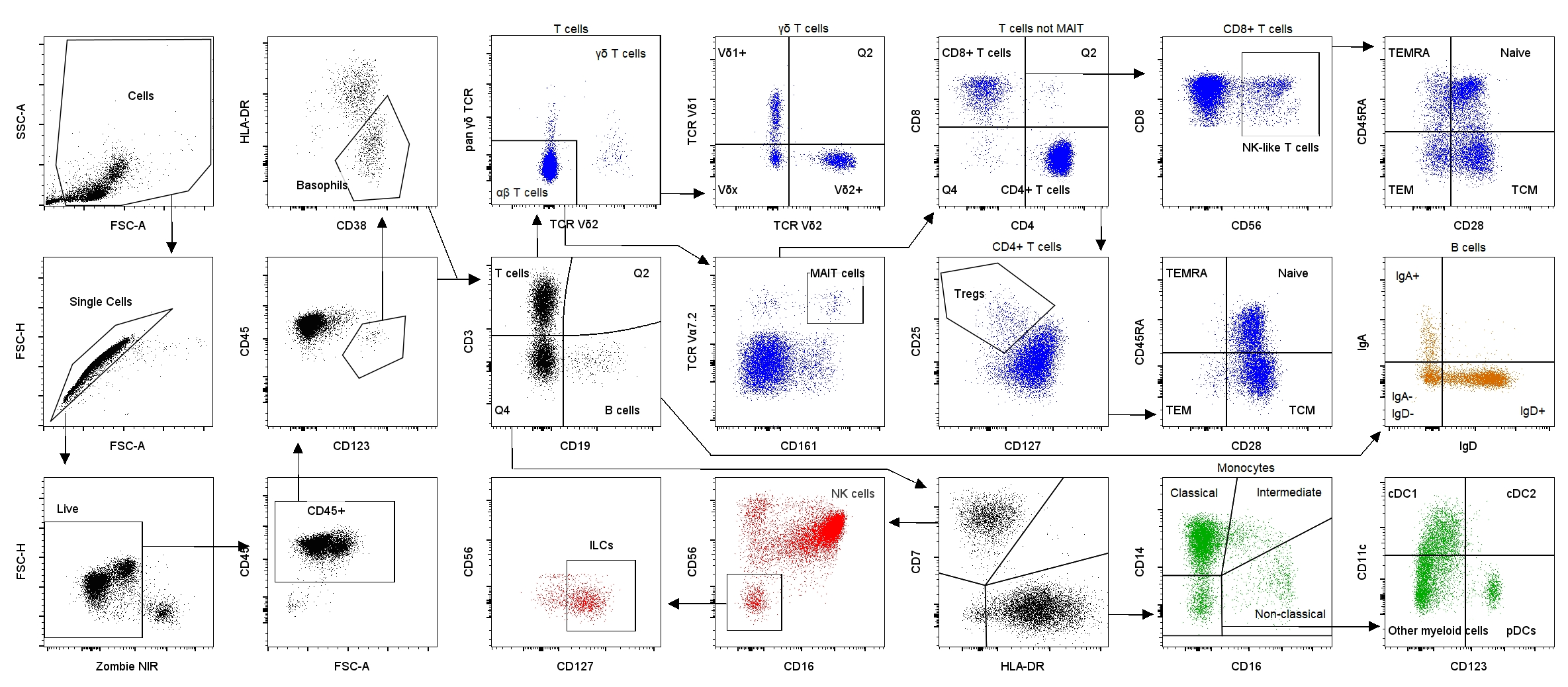

### Supplemental Figure 2

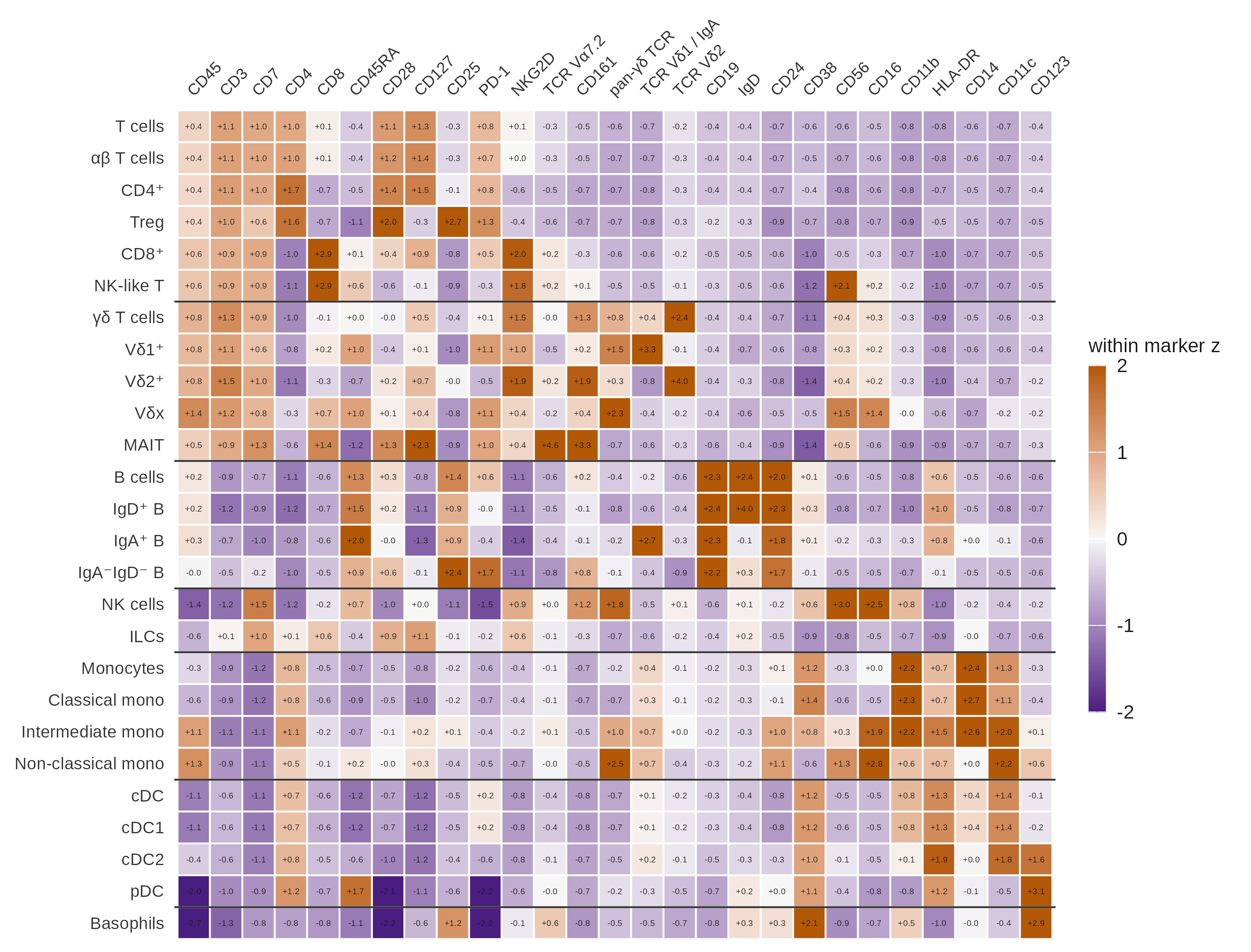

### Supplemental Figure 3

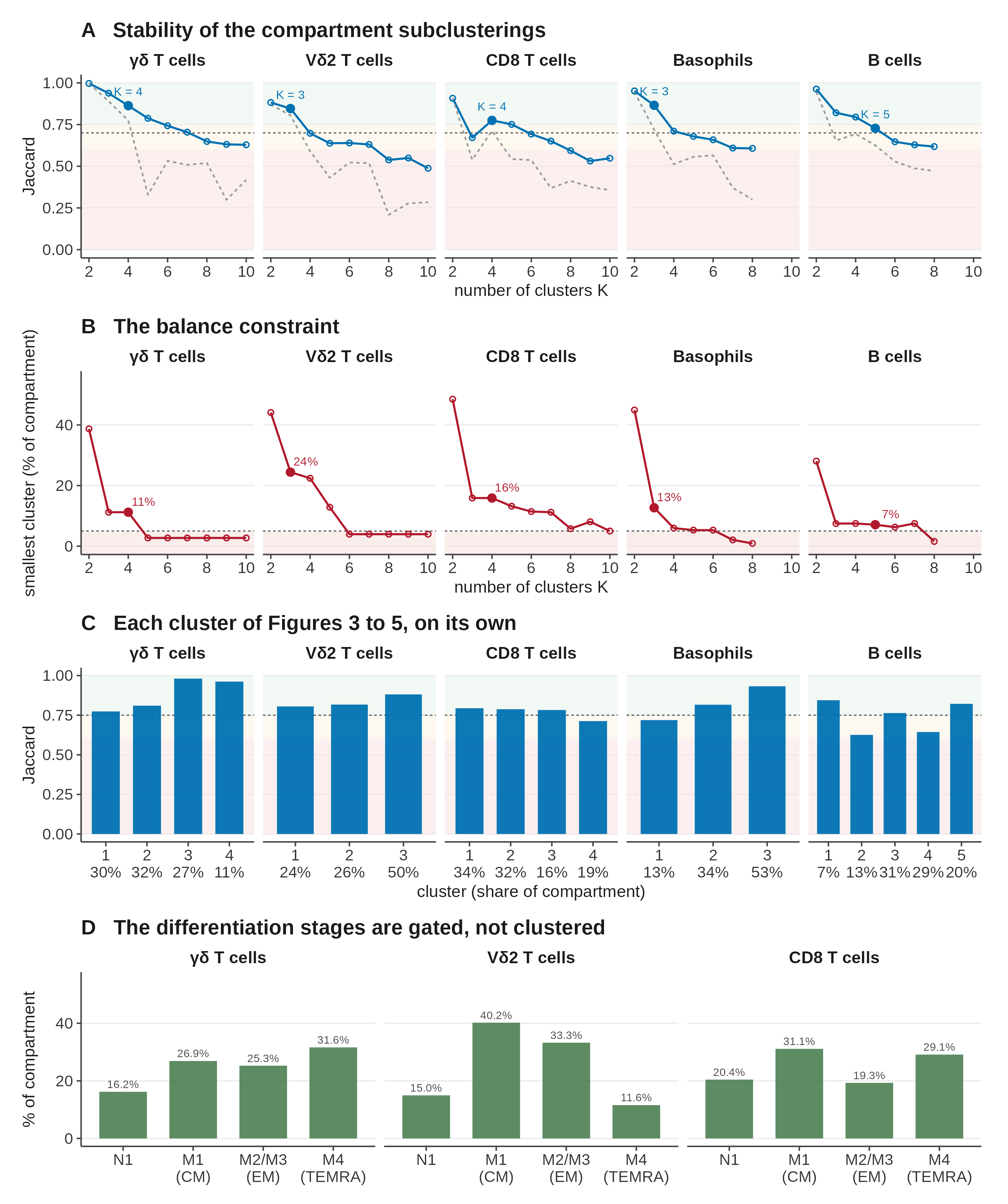

### Supplemental Figure 4

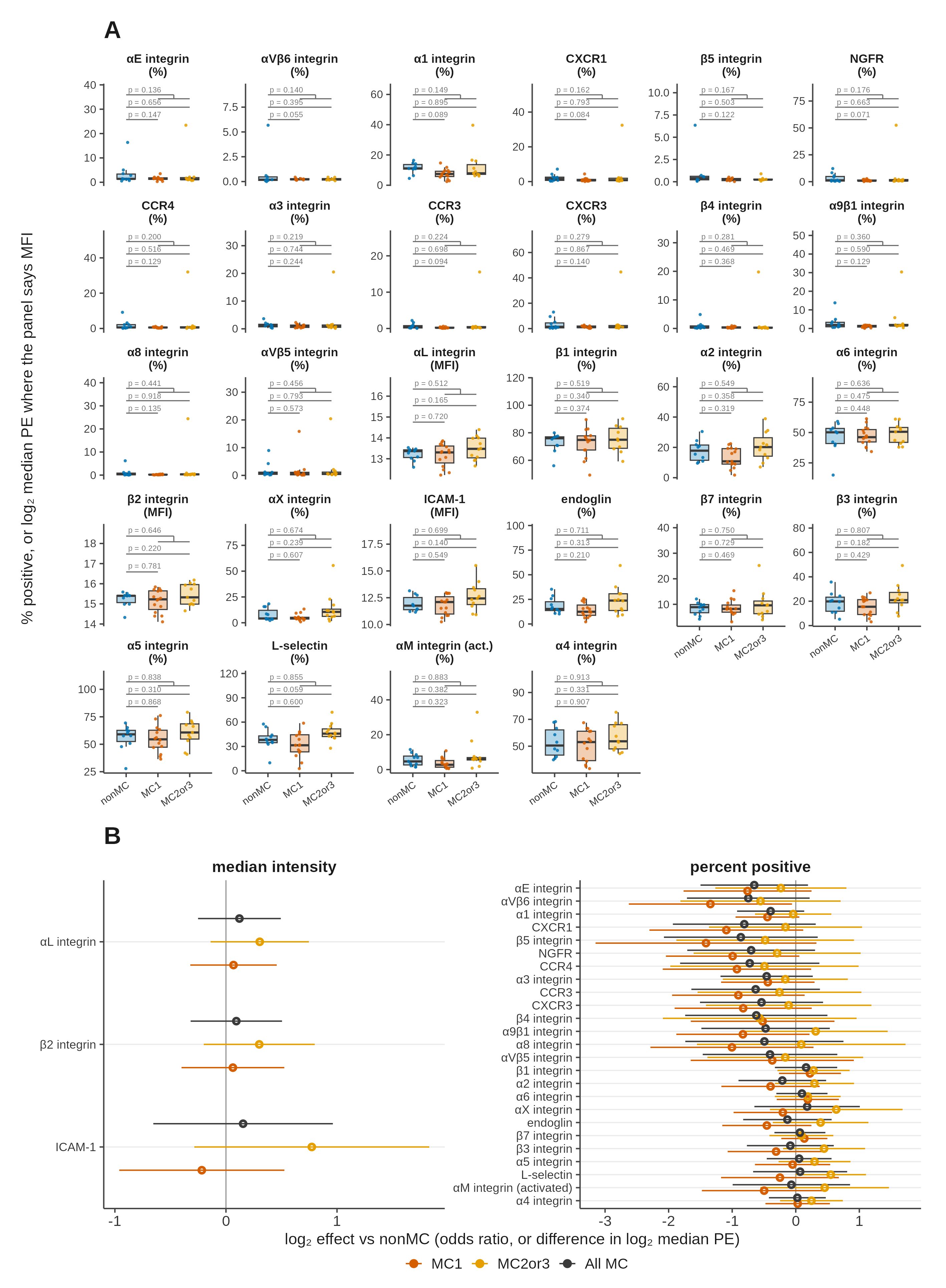
